# The relationship between shared and differentiating genetic liability for schizophrenia and bipolar disorder and cognition and educational attainment in the UK Biobank

**DOI:** 10.1101/2025.02.13.25322048

**Authors:** Alexander L Richards, Eilidh Fenner, Nicholas E Clifton, Darren Cameron, Claire E Tume, Nicholas J Bray, Sophie E Legge, James TR Walters, Peter A Holmans, Michael C O’Donovan, Michael J Owen

## Abstract

**Importance:** Further understanding how genetic liabilities to schizophrenia (SZ) and bipolar disorder (BD) are related to cognition and educational attainment (EA) indicates important differences between these conditions that are relevant to future research and interventions.

**Objective:** To determine how fractions of genetic liability that are shared between, and differentiate, SZ and BD, are associated with cognition and EA and characterise their biology using gene-set enrichment analysis.

Design, setting and participants: Fractions of liability were derived using Genomic Structural Equation Modelling (gSEM) on genome-wide association studies of SZ and BD. Polygenic risk scores (PRS) representing each fraction were tested for association with two measures of cognition - a general cognitive factor *g* (n=93451) and fluid intelligence (FI, n=160465) and a measure of EA (n=354609) in the UK Biobank, excluding individuals with SZ, BD or a psychotic disorder. MAGMA and partitioned LDSC were used to examine the shared and differentiating fractions for enrichment in genes with high expression specificity for cell types, functional categories and developmental stages.

Main outcomes and measures: PRS representing the shared and differentiating fractions were tested for association with FI, *g* and EA. Mean -log10 p-value across both MAGMA and partitioned LDSC results was used as a metric of enrichment.

**Results:** The shared fraction was associated with poorer cognition (FI beta -0.079 p=5.57e-85; *g* beta -0.079, p =7.51e-51) but higher EA (beta 0.016, p=5.08e-07). The SZ differentiating fraction (SZ_diff_) was associated with poorer cognition (FI beta -0.027, p=6.34e-24; *g* beta -0.009, p=3.74e-3) and lower EA (EA beta -0.049, p=1.84e-58). The BD differentiating fraction (BD_diff_) was associated with better cognition and higher EA, the effects being of the same magnitude as SZ_diff_ but of opposite sign. Adjusting for cognitive function, the effects of SZ_diff_ PRS on EA were attenuated but remained significantly (adjusted for FI, beta -0.025, p = 1.28e-6; adjusted for *g,* beta *-*0.032, p = 8.87e-6). The differentiating fraction was enriched for genes specifically expressed in young adulthood (20-30 years) and mid adulthood (30-60 years), but not in earlier developmental stages. It was also enriched in murine pyramidal CA1 cells and striatal medium spiny neurones.

Conclusions and relevance: Our findings partly explain the greater cognitive impairments and stronger negative genetic correlation with intelligence in SZ compared to BD. This may reflect neurodevelopmental processes indexed by cognitive function that are more prominent in SZ than BD, although the relatively modest effect of SZ_diff_ on cognition is consistent with studies suggesting that inherited genetic variation is not the major determinant of cognitive impairment in SZ. Despite negative effects on cognition, the shared fraction is weakly associated with better EA, suggesting that this is the result of influences on noncognitive traits. In contrast, the SZ_diff_ fraction is enriched for alleles that confer risk to poorer EA through both cognitive and non-cognitive mechanisms, which has implications for interventions. The differentiating fraction was enriched for genes with relatively high expression specificity in early and mid-adulthood, corresponding to the typical age at onset of psychotic symptoms, a clinical feature that has previously been associated with this fraction of liability.

## Introduction

Schizophrenia (SZ) and bipolar disorder (BD) are highly heritable polygenic psychiatric disorders^1^. Although they are classified as distinct entities in the major diagnostic systems (ICD-11, and DSM-5)^2,3^, their clinical features substantially overlap^4^ and there is strong evidence for shared risk factors including genetic liability, where estimates of the genetic correlation between the two are typically in the range of about 0.7^5^. The evidence for clinical and aetiological overlap between the two disorders is consistent with the proposition that SZ and BD occupy different, but nevertheless overlapping, positions on several dimensions of psychopathology rather than as fully independent categories of disorder^6^. This hypothesis is further supported by studies that have shown that risk alleles that influence major dimensions of symptomatology (e.g. psychosis, depression, mania) are in part distinct, and moreover, that the same alleles influence those dimensions across diagnoses^7–10^.

Cognitive impairment is also a feature of SZ and BD. It is typically more severe in the former than in the latter^11–13^ and involves most aspects of cognitive function^14^ including IQ. In BD, while the deficits are milder, they are qualitatively similar^15–17^. While consistent with a dimensional view of SZ and BD, these findings suggest that there may be a set of pathogenic processes that are manifest by cognitive impairment and which are more prominent in those diagnosed with SZ rather than BD. Moreover, while cognitive impairment is not a core diagnostic criterion for either SZ or BD, it is strongly associated with functional outcomes in areas such as work and independent living^14^ and therefore of considerable importance to people with these disorders, their families and to people involved in their clinical care. Understanding the aetiology of cognitive impairment in SZ and BD is therefore important topic as this may point to potential therapeutic interventions that can improve outcomes^18^ or potentially even prevent illness onset should low cognitive ability be established to be a causal risk factor for the disorders.

It has been postulated that cognitive impairment seen in SZ reflects an underlying perturbation of neurodevelopment, and that this is more prominent in SZ than in BD^19,20^. Predictions that follow from this hypothesis are that the genetic fraction that differentiates SZ from BD will include variants associated with cognition, and that these variants will be enriched in genes whose expression characterises early brain development. Moreover, the alleles at those sites that are associated with poorer cognition will be those that preferentially increase liability to SZ over BD.

Here we test those predictions. We applied Genomic SEM (gSEM)^21^ to large genome wide studies of SZ and BD to isolate the fraction of common variant liability that differentiates between the two disorders as well as the fraction of liability that they share. We then used genetic correlation and polygenic risk score (PRS) analyses to examine the relationships between these fractions and common genetic influences on cognition. Aiming also to illuminate possible biological processes that might mediate these effects, we sought to identify gene sets and cell populations, and developmental time points, that are enriched for the differentiating fraction of liability.

We also sought to better understand the relationships between genetic liability to SZ and BD, cognitive ability and educational attainment (EA). Our motivation here was two-fold. First, in the absence of very large case samples in which direct measures of cognitive ability, are available, EA is often used as a proxy measure of cognitive ability in genomic studies. Second, some^22–25^ though not all^26–28^ studies have reported the surprising finding that genetic liability to schizophrenia shows a small positive association with genetic liability for better educational outcomes despite the robust evidence for association between the disorder and both lower cognitive ability and poorer educational outcomes^29^.

## Methods

### Genomic SEM

The GWAS summary statistics for gSEM were from studies of SZ and BD conducted by the Psychiatric Genomics Consortium (PGC)^30,31^ (Supplementary Table S1). The BD GWAS included only participants of European ancestry, so we similarly restricted the SZ GWAS summary statistics to those of European ancestry to avoid ancestry related differences in allele frequency and patterns of linkage disequilibrium confounding our estimates of genomic differences between the disorders^7^.

Single nucleotide polymorphisms (SNPs) were included in the analyses if they had a minor allele frequency greater than 1% in HapMap 3 reference set^32^, were present in both source GWASs, and had an imputation score of at least 0.7. Variants within the extended MHC were excluded (chromosome 6; 25-35 Mb) as it is difficult to allow for the complex LD structure in the region. We retained 7,334,582 SNPs for analysis. We used gSEM to apply a common factor model to the summary statistics from the GWAS (see Supplementary Methods). gSEM was run in R (The R Foundation, version 4.0.3) using the GenomicSEM package^21^. gSEM estimates and corrects for sample overlap among the input GWAS^33^. For each SNP, the loading on the common factor was extracted to produce a statistic corresponding to the effect of that variant that is shared between SZ and BD. We then applied a model where we extracted the loading of each SNP on the residual variance from each input GWAS that was not explained by the common factor (see Supplementary Methods) so that the residual effect sizes for each SNP indexes how much it influences the probability of having one phenotype over the other. The SZ differentiating fraction (SZ_diff_) denotes effects signed such that beta above zero indicates an allele that increases the probability of SZ over BD, while those below zero indicate the reverse. Conversely, we refer to the bipolar differentiating fraction (BD_diff_) to denote effects signed such that beta above zero indicates an allele that increases the probability of BD over SZ, while those below zero indicate the reverse. Note that in the present study, as there are only two phenotypes in the model, SZ_diff_ and BD_diff_ are perfectly negatively correlated.

SNP-based heritabilities (SNPh2) and genetic correlations were calculated using linkage disequilibrium score regression^33,34^. We report heritability on the observed scale because the population prevalences of the latent constructs underlying gSEM fractions are unknown, precluding conversion to a liability scale.

### Cognitive and Education datasets

We tested for genetic correlations between the input GWAS and gSEM fractions and IQ and educational attainment (EA) from publicly available summary statistics^35,36^. We also used a PRS approach^37^ to test for association between gSEM fractions of liability and measures of cognition and EA in the UK Biobank, a UK prospective volunteer study of around 500,000 participants aged 40-69 at the time of recruitment (www.ukbiobank.ac.uk). The North-West Multi-Centre Ethics Committee granted ethical approval to UK Biobank, and all participants provided written informed consent. This study was conducted under UK Biobank project number *13310*.

### Genotyping in UK Biobank

Exclusion criteria for genetic variants in the UK Biobank data were: genotyping rate < 0.95, minor allele frequency < 0.01, Hardy-Weinberg equilibrium (HWE) p-value < 1e-6 (using the ‘midp’ and ‘keep-fewhet’ options), imputation INFO score < 0.9. Individuals were excluded if they had SNP missingness > 0.05.

### Phenotyping in UK Biobank

We excluded people with a diagnosis of bipolar disorder, schizophrenia or a psychotic disorder based on primary care data, hospital inpatient data, death register records, or self-report.^38^

Cognitive assessments in the UK Biobank were brief and unsupervised. Individual cognitive test scores from this dataset vary in reliability and stability^39,40^, but performance between different cognitive tests is correlated^40,41^ which enables a more robust measure of generalised cognition (*g*) to be derived from a principal component analysis of multiple cognitive tests^39^. Our approach to deriving a measure of *g* and the rationale for the inclusion/exclusion of individual cognitive tests are provided elsewhere^42^. Briefly, *g* was derived as the first principal component from analysis of: numeric memory (from the first online cognitive test battery); reaction time (from baseline assessment); pairs matching (from baseline assessment); and trail making test B (from the first online cognitive test battery). *g* was only available for participants who had completed all four tests (n = 94956, Supplementary Tables S2)*. g* was standardised and outliers were removed (*g* score > 4 or < -4). The UK Biobank also has data available on a subset of participants for a measure of fluid intelligence (FI). FI was measured at either the initial assessment or one of two subsequent assessments. FI was standardised, then if it was measured at multiple timepoints, we took the mean value. FI was not included in our estimate of *g*, allowing us to test the relationship between the gSEM fractions and FI as a check of robustness of our findings in relation to *g*.

The Biobank data included a variable representing level of education achieved by participants. As in our previous publication^26^, we used these data to derive an ordinal measure representing the highest educational level the participant had achieved (1=no qualifications, 2=NVQ/HND/HNC or equivalent, 3=CSE or equivalent, 4=O-level/GCSE or equivalent, 5=A/AS-levels or equivalent, 6=college or university degree). Professional qualifications were not considered, as the level of academic achievement they represent varies by profession.

### Polygenic Risk Score (PRS) Analyses

PRS were derived as described^37^. Clumping was performed on imputed best-estimate genotypes using PLINK (maximum r2=0.2; window=500 kb; minimum minor allele frequency=0.1; minimum Info score=0.7). Variants within the extended MHC were excluded (chromosome 6 from 25 MB to 35 MB). As optimal P value thresholds for inclusion of alleles in the gSEM-derived PRS are unknown and no large independent samples are available to derive them, we performed PRS analysis without P value thresholding^7^.

Association tests were adjusted for the first 10 population principal components, sex, age at interview, age at interview squared and genotyping platform. Educational attainment analyses were adjusted for birth before 1950 to reflect changes in the educational grading system in the UK^43^. All PRS variables were standardized before analysis using the scale() function in R. We tested PRS for association with *g* (93541 participants, Supplementary Table S3) and FI (160465 participants) using linear regression, reporting beta and P values for the PRS term in the regression model. PRS were tested for association with the ordinal measure corresponding to the highest educational level achieved using ordinal regression (354609 participants). All P values were 2-tailed.

### Calculating Gene Expression Specificity Across Cell Populations

Gene expression specificity scores were obtained from four independent datasets: two single-nucleus RNA-Seq studies in human and two single-cell RNA-Seq studies in mouse. These included 91 cell populations across five regions of the second-trimester fetal brain^44^, 84 populations from the human prefrontal cortex spanning gestation to adulthood^45,46^, 24 populations from the mouse brain^47^, and 15 populations from adult human hippocampus and prefrontal cortex^48^. Further details are provided in Supplementary Tables S5. Gene expression specificity scores were calculated by dividing each gene’s normalized unique molecular identifier (UMI) count in a given cell type by the sum of that gene’s expression across all cell types. Genes mapping to the extended MHC region or chromosome X were excluded. As with the developmental stage analyses, the top 10% of genes with the highest specificity scores for each cell type were selected.

### Evaluating Genetic Liability Enrichment in Stage- and Cell-Specific Gene Sets

To evaluate whether the genetic liability captured by the gSEM “shared” and “differentiating” fractions of schizophrenia and bipolar disorder is associated with genes showing specificity for developmental stages or cell types, two independent analyses were performed: a gene-based association test using MAGMA (v1.10)^49^ and a partitioned heritability analysis using LD score regression (LDSC, v1.2)^33,50^.

For the MAGMA analyses, SNP-level p-values were aggregated into gene-level statistics using a snp-wise=mean model, with a 35 kb upstream and 10 kb downstream window applied to each gene’s coding sequence. One-sided tests assessed each gene set for enrichment for associations in the gSEM shared and differentiating fractions. One-sided competitive p-values for each gene set werer extracted as the primary test statistics. For partitioned LDSC, we followed Finucane et al. (2018) by applying a 100 kb window around each gene and incorporating version 1.2 of the baseline LD annotations. The one-sided coefficient z-score p-value was extracted as a measure of heritability enrichment in the gSEM shared and differentiating fractions in each gene set. For comparison, MAGMA and partitioned LDSC enrichment analyses were also performed on the SZ and BD source GWAS.

### Quantifying Gene Expression Specificity Across Developmental Stages

Transcriptomic data from the dorsolateral prefrontal cortex (DLPFC) and hippocampus were obtained from the BrainSeq Phase II database^51^. This consisted of 300 DLPFC and 314 hippocampal samples from 374 individuals, ranging from 12 post-conceptual weeks to 84 years (further details in Supplementary Table S4). Samples were divided into developmental stages defined in Supplementary Table S5. Raw read counts were normalized using the trimmed mean of M-values (TMM) method^52^. For each gene, expression values were regressed against a single developmental stage, modelled as a binary variable indicating whether a sample belonged to the stage under consideration or not. Brain region, sex, and genetic ancestry, represented by the first five principal components of genotype, were included as covariates, and within-individual correlation was accounted for using the duplicateCorrelation function in the limma R package. From each model, the t-statistic for the developmental stage term was extracted for every gene, serving as a measure of developmental stage expression specificity relative to all other stages^53,54^. The top 10% stage-specific genes, ranked by their t-statistics, were selected to define stage-specific gene sets.

### Gene Ontology Enrichment

We also included a gene set derived from 7,296 Gene Ontology (GO) categories in the MAGMA analyses. Gene Ontology categories were downloaded from the Gene Ontology Consortium^55^ (geneontology.org, November 9, 2020). In the curation process, low-confidence gene-category relationships, such as those inferred from electronic annotation (IEA), non-traceable author statements (NAS), or reviewed computational analysis (RCA), were excluded, along with obsolete categories^56^. After curation, MAGMA was run as described above on the schizophrenia and bipolar disorder differentiating and shared fractions, and the source GWASs.

### Results

### Heritability and Genetic Correlations

Table 1 reports the estimated heritability from the source GWAS datasets as well as the gSEM fractions. We report heritability on the observed scale as the absence of population prevalence data for the latent gSEM constructs preclude deriving values on the liability scale. As expected, given the strong genetic correlation between schizophrenia and bipolar disorder, for gSEM, the majority of heritability was attributable to the fraction that is shared between the disorders.

**Table 1.** Heritability of gSEM fractions and SZ/BD GWAS.

| **Input GWAS** | **Heritability (observed scale)** | **S.E heritability** |
| --- | --- | --- |
| Schizophrenia | 0.35 | 0.01 |
| Bipolar Disorder | 0.28 | 0.01 |
| Shared | 0.26 | 0.01 |
| Differentiating | 0.14 | 0.01 |

Genetic Correlations are given in table 2. Liability to schizophrenia was negatively correlated with that for (higher) IQ (rg –0.22) as was, less strongly, liability to BD (rg –0.07). Liability to higher IQ was correlated with liability to higher EA (rg=0.74). Despite negative correlations with IQ, liability to schizophrenia was not significantly associated with EA (rg 0.02) while liability to BD was significantly associated with liability to higher EA (rg 0.13). Almost identical estimates of genetic correlation between genetic liabilities to the disorders and both intelligence and EA have been reported using a different methodology^27^.

**Table 2.** Genetic correlation for schizophrenia^31^ and disorder GWASs^30^, gSEM shared and schizophrenia differentiating (SZ_Diff_) fractions (derived in the present study) and published IQ^35^ and Education Attainment (EA)^36^ GWASs from general population samples. Correlations were calculated using LDSC. Genetic correlation (rg) values are below the diagonal with standard errors in brackets. Genetic correlation p-values are given above diagonal.

|  | Schizophrenia | Bipolar Disorder | Shared | SZ<sub>Diff</sub> | IQ | EA |
| --- | --- | --- | --- | --- | --- | --- |
| Schizophrenia |  | <1E-300 | <1E-300 | 1.86E-26 | 1.80E-25 | 0.18 |
| Bipolar Disorder | 0.68 (0.017) |  | <1E-300 | 1.08E-98 | 0.0006 | 4.30E-12 |
| Shared | 0.94 (0.003) | 0.90 (0.006) |  | 0.015 | 9.65E-19 | 2.02E-06 |
| SZ<sub>Diff</sub> | 0.29 (0.027) | -0.49 (0.023) | -0.06 (0.024) |  | 1.82E-09 | 1.71E-06 |
| IQ | -0.22 (0.027) | -0.07 (0.022) | -0.17 (0.019) | -0.16 (0.027) |  | <1E-300 |
| EA | 0.02 (0.025) | 0.13 (0.018) | 0.07 (0.016) | -0.12 (0.025) | 0.74 (0.01) |  |

The shared fraction of liability also showed discordant effects, being negatively correlated with liability to higher intelligence (rg -0.17) but positively correlated with liability to higher EA (0.07). In contrast, the SZ_diff_ fraction showed congruent effects, being associated with both lower cognition (rg -0.16) and lower educational attainment (rg -0.12). From the perspective of BD, this is the equivalent of the BD_diff_ fraction being correlated with better cognitive ability and with better EA.

### Polygenic Risk Score Analyses of Cognition in UK Biobank

The results of polygenic risk score (PRS) analyses of cognition are given in Figure 1 and the detailed results in Supplementary Table S6. The SZ, BD, and the shared gSEM fractions were negatively associated with *g*; schizophrenia beta -0.075, p=4.16e-51; BD beta -0.035, p=9.05e-17; shared beta -0.079, p=7.51e-51. The fraction of liability that differentiates between schizophrenia and bipolar disorder was weakly associated with lower *g* (SZ_diff_ beta -0.009, p=3.74e-3), or, reciprocally, from the perspective of BD, alleles that differentiate BD from SZ were weakly associated with higher *g*. The pattern of associations with FI were similar to those obtained with *g*, but with a stronger effect for the differentiating fraction (Figure 1, Supplementary Table S5). Note that the relative magnitudes of effects for different PRS are not meaningful as they are dependent not only with the degree of shared genetic liability with the cognitive measures, they are also dependent on the power of relevant input GWAS.

**Figure 1.**
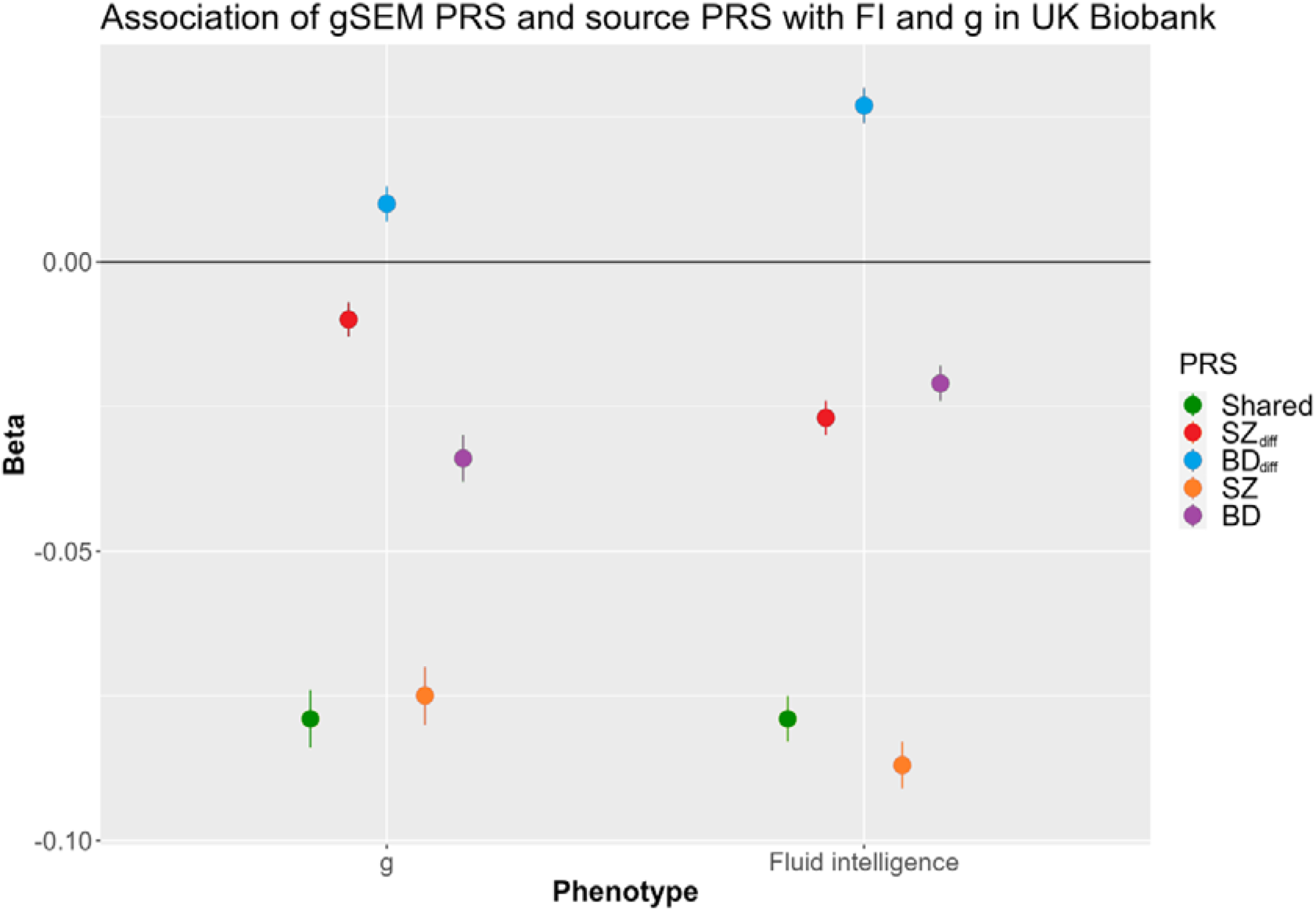
Association of gSEM fractions and source GWAS PRS with *g* and fluid intelligence in UK Biobank (fluid intelligence N= 160465; *g* N= 93541). Point estimates for beta with standard errors are given. Negative betas indicate higher liability to the relevant trait is associated with poorer cognition.

### Polygenic Risk Score Analyses of Educational attainment in UK Biobank

The results of polygenic score analyses of EA are given in figure 2 with full detail in Supplementary Table S7. Schizophrenia PRS was associated with lower EA (beta -0.013, p=2.46e-5) while the BD PRS was associated with higher EA (beta 0.043, p=9.07e-44). Consistent with genetic correlation analysis, the gSEM shared fraction PRS was associated with higher EA (beta 0.016, p=5.08e-7) while the SZ_diff_ fraction was associated with lower EA (beta -0.049, p=1.84e-58). Reciprocally, the PRS representing alleles that favour the development of BD rather than SZ at those sites that differentiate the two disorders were associated with better educational outcomes.

**Figure 2.**
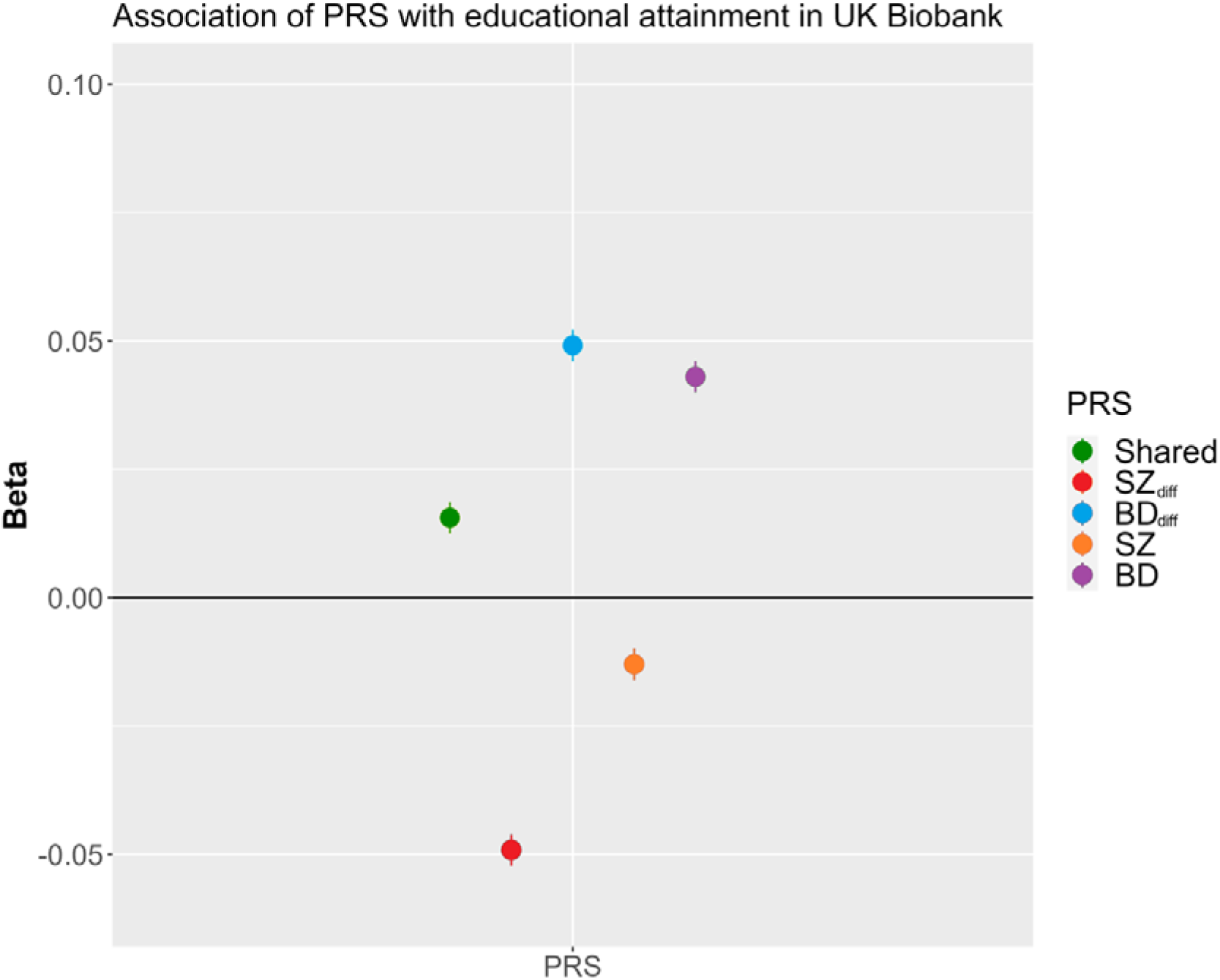
Association of gSEM PRS and source PRS with Educational Attainment (EA) in UK Biobank. Point estimates for beta with standard errors are given. Negative beta values indicate higher liability to the relevant trait is associated with lower EA.

### Cognitive and non-cognitive effects on Education

The discordant effects of the shared liability fraction on cognition and EA as indexed by both genetic correlation and PRS analyses allow us to infer that the shared fraction of liability is enriched for alleles that promote higher educational outcomes through non-cognitive mechanisms. In contrast, the concordant effects on cognition and education of the differentiating genetic fraction suggest that alleles that favour the development of schizophrenia over BD likely exert effects on EA through negative effects on cognition, but it is also possible they influence the non-cognitive aspects of that trait. To explore that possibility, for our primary analysis, we re-tested the effects of the PRS for SZ_diff_ on EA covarying for fluid intelligence rather than *g* as the former is better powered given more data are available. The effects of SZ_diff_ PRS were attenuated but remained significantly associated with poorer educational attainment (unadjusted analysis of EA on subset of samples with FI data: beta -0.042, se=0.005, p=2.55e-17; adjusted for FI: beta -0.025, se=0.005, p=1.28e-6) indicating that this fraction is enriched for alleles that promote poor EA outcomes through non-cognitive as well as cognitive effects. The association with EA also remained after adjusting for *g* in the smaller sample size in which it could be derived, although the degree of attenuation was negligible consistent with the weak association between SZ_diff_ and *g* (unadjusted analysis on subset of samples with *g* data: beta -0.035, se=0.007, p=1.02e-6; analysis adjusted for *g:* beta -0.032, se=0.007, p=8.87e-6).

### Enrichment analyses

We did not find consistent evidence (i.e. across both methods of analysis) for enrichment of associations in the shared fraction for genes with high specificity for any developmental stage (Fig 3, Supplementary Table 8). However, genes with high expression specificity for young (age 20-30 years) and mid (age 30-60 ranges) adulthood were significantly enriched for associations that differentiate between SZ and BD. Individual gene-wide p values for all genes including annotation for high expression specificity with respect to young and mid-adulthood are provided in Supplementary Table S13. Genes with high expression specificity in these two adult gene sets only modestly overlap (Supplementary Table 13). Moreover, of 20 genes with significant differential fraction MAGMA (P<2e-6) associations in the top deciles for either of these two developmental stages, only 4 were in the top decile for both of these stages of adulthood (Supplementary table 13). Limited overlap was seen at more relaxed significance thresholds indicating the enrichments at each of these developmental stages represent essentially independent signals (Supplementary Figure S1). The BD GWAS showed stronger enrichment for associations in these same sets of genes compared with the source SZ GWAS (Fig 3) suggesting this differentiating association enrichment is driven by alleles that increase liability to BD rather than those that that increase liability to SZ. Note that the SZ GWAS showed stronger evidence than the BD GWAS for enrichment for associations in genes with high expression specificity for late-midfetal and early infancy stages but this was not accompanied by enrichments for any of the gSEM fractions.

**Figure 3.**
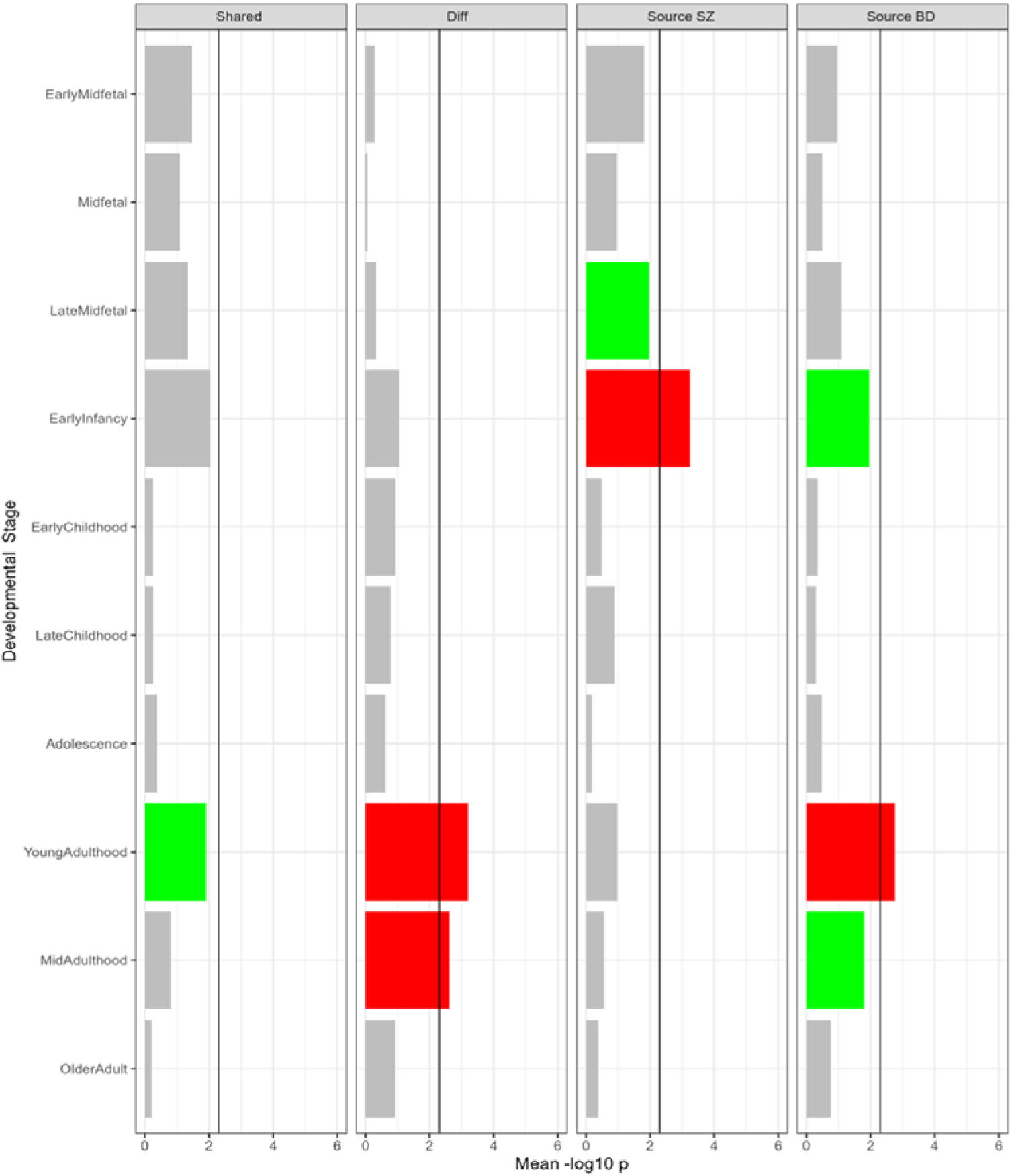
Enrichment of fractions of liability in genes with high specificity for developmental stages. Mean -log10 p indicates mean of the -log10 significance level for MAGMA and partitioned LDSC (pLDSC) enrichment tests. The black line represents the corrected significance threshold (Bonferroni corrected for 10 developmental stages). Red bars indicate significance threshold reached in both MAGMA and pLDSC, green in MAGMA only, and blue in pLDSC only. EarlyMidfetal samples are between 10 post-conception weeks (pcw) and 16 pcw, Midfetal between 16 pcw and 17 pcw, LateMidfetal from between 17 pcw and 24 pcw, EarlyInfancy between birth and 6 months of age, EarlyChildhood between 1 and 6 years, LateChildhood between 6 and 13 years, Adolescence between 13 and 20 years, YoungAdulthood between 20 and 30 years, MidAdulthood between 30 and 60 years, and OlderAdult over 60 years.

Details of the cellular enrichments are provided (Supplementary Tables S9-S12, Supplementary Figures S2 to S6). We found no consistent evidence that the differentiating fraction of liability was enriched for genes with high single nuclear expression specificity for cell populations in human second trimester fetal brain^44^ (Supplementary Table S9), human prefrontal cortex at multiple time points from gestation to adulthood^45^ (Supplementary Table S12), or for cell types from adult human prefrontal cortex and hippocampus^48^ (Supplementary Table S11). However, in cell types from mouse brain^47^ (Supplementary Table S10), we did see evidence that the differentiating fraction was enriched for genes with high single cell expression specificity for excitatory glutamatergic pyramidal neurones from the CA1 region of the hippocampus, and from GABAergic inhibitory medium spiny neurones (MSN) of the striatum. Genes with high expression specificity for the same cell types were also enriched for signal in the shared fraction of liability and in each of the source GWAS. There was very little relationship between the gene-wide differentiating and the shared association test statistics for genes with high expression specificity for the MSN and pyrCA1 gene sets (MSN r = 0.17, pyrCA1 r=0.018) suggesting that different genes drive the shared and differentiating liability in these cell types.

We found 29 genes with significant (P<2e-6) MAGMA associations in at least one of the MRN or pyrCA1 cell types, 19 of which showed no evidence (P>0.05) for association in the differentiating fraction. Genes with specificity for the association in the shared fraction included many well-known candidates including calcium channels *CACNA1C* and *CACNB2,* dopamine receptor *DRD2*, glutamatergic receptor subunit *GRIN2A*, and the cholinergic receptor subunit *CHRM4*. Fewer genes in these two gene sets (N=9) were associated in the differentiating fraction, 5 of which showed no evidence in the shared fraction. Individual gene-wide p values for all genes including annotation for high expression specificity with respect to CA1 and MSN gene-sets are provided in Supplementary Table S13.

Analyses of gene ontology (Supplementary Table S14) revealed the shared fraction of liability highlighted similar biological processes and molecular functions as the source GWAS of SZ and of BD, though more categories were significant corrected for 7296 categories) in the shared component (N=58 in shared, N= 38 in SZ, N= 11 in BD). Moreover 36/38 of the categories that were significant in SZ and 10/11 of those that were significant in BD were also significantly enriched in the shared component, and only 1 (GO:0098797 plasma membrane protein complex, significant in BD) of these 3 categories did not have moderate evidence (nominal P<0.001) in the shared category. Gene Ontology analyses of the differentiating fraction identified no significant findings (Supplementary Table S14).

## Discussion

The primary aim of this study was to clarify the relationship between cognitive ability and the fraction of liability that is shared between SZ and BD and that fraction which differs between the two disorders. Our main hypothesis was that the fraction that confers differential liability to SZ and BD will contain variants associated with cognition and that the specific alleles at those sites that are associated with poorer cognition will be those that preferentially increase liability to schizophrenia over bipolar disorder. We also sought to obtain insights into the possible biological processes, cell types, and developmental stages through which differential liability is mediated, predicting the associations for the differential component would be enriched among genes showing high expression specificity for early neurodevelopment.

Our findings are consistent with the primary hypothesis that genetic liability that is relatively specific to SZ is associated with poorer cognitive performance in the general population while, conversely, genetic liability that is relatively specific to BD is associated with higher cognitive ability. We also showed that the fraction that is shared between the two disorders is associated with poorer cognition. This pattern of findings is in accord with the observations that both disorders are associated to a degree with cognitive impairment and suggests that this reflects to some extent common alleles that are shared across the disorders and which confer risk both to mental illness and poorer cognitive function in the general population. Our finding that the SZ_diff_ fraction was associated with poorer cognition, and conversely the BD_diff_ fraction was associated with better cognition, provides at least a partial explanation for the greater impairments in cognitive performance and stronger negative genetic correlation with intelligence in SZ compared to BD^30,57^. It also supports the notion that the more severe cognitive impairment seen in schizophrenia reflects in part neurodevelopmental processes and mechanisms related to the development of cognitive function that are more prominent in SZ than BD^19,58^. However, the relatively modest effect of SZ_diff_ on cognition is in line with a substantial body of evidence that inherited genetic variation is not the major determinant of cognitive impairments in SZ and that these are more strongly associated with non-inherited factors indexed by cognitive performance that deviates significantly from familial expectations^29^. There is also evidence that these non-familial factors play a greater role in SZ than BD^59^ and, together with our findings, this suggests that these may be more important than common genetic variation in the greater cognitive impairment seen in SZ than BD.

As far as we are aware, ours is the first study to compare the relationships between shared and specific fractions of genetic liability to SZ and BD with direct measures of cognitive function. Our results are, however, consistent with those from studies using different methods to compare genetic liability to SZ with that for BD. A study based on conditional false discovery rate (condFDR) analysis reported that both SZ and BD showed substantial genetic overlaps with cognitive function^60^. It also found that among individual alleles that showed fairly strong evidence (FDR<0.01) for joint association with schizophrenia and intelligence (N=75), most (81%) were associated with lower cognitive performance while among BD alleles jointly associated with intelligence (n=12), most (75%) were associated with better cognitive performance. A subsequent study by the same group^27^ using a bivariate causal mixture model (MiXeR) showed extensive genetic overlap between the variant sites that influence each of intelligence and EA and those that confer liability to BD and SZ. They also showed low to moderate disorder-trait genetic correlations, as we do here and as have others^30,57^. As genetic correlations are genome-wide measures of similarity of allelic effect directions and sizes between traits, the modest genetic correlations in the face of extensive overlapping sites implies that liabilities to SZ and to BD include a mix of alleles with different directions of effect on intelligence and EA. Further analyses using LAVA^61^ also showed prominent mixed directions of effect between BD, schizophrenia, and cognitive traits.

A secondary aim of our study was to examine the relationships between the genetic fractions of liabilities to SZ and BD and liability to EA as well measured EA performance. The fraction of liability that is shared between SZ and BD was weakly but significantly genetically correlated with liability to higher EA (table 1) and also with better measured EA performance (figure 2) while the SZ_diff_ fraction was negatively correlated with liability to higher EA and was associated with poorer EA performance. Schizophrenia liability therefore includes a greater proportion of risk alleles that negatively influence educational outcomes than does liability to BD, which explains why overall liability to BD is associated with better educational outcome and overall liability to SZ is associated with poorer educational outcomes despite the high genetic correlation between the two disorders.

Our study also extends previous work on the relationships between the cognitive and non-cognitive components of educational attainment and the shared and specific fractions of liability to SZ and BD^62,63,21^ by incorporating direct measures of cognition as well as estimates of educational attainment. The counter-intuitive observation that, while shared liability is associated with poorer cognition (figure 1), it is also associated with higher EA (figure 2) implies that most of the heritability conferred by this fraction of liability comes from alleles that are associated with *noncognitive* traits that promote EA. In contrast, the observation that SZ_diff_ is associated with poorer cognition <u>and</u> with poorer EA suggests that this fraction of liability includes alleles with *negative* effects on the *cognitive* fraction of EA. This assertion is supported by the attenuation in the association between SZ_diff_ and EA seen after conditioning on cognitive ability, particularly FI. However, given that SZ_diff_ fraction remained associated with poor EA after conditioning, it must also include alleles that are associated with *noncognitive* traits that promote *lower* EA. Nevertheless, given that overall liability to SZ shows little (based on PRS) to no (based on genetic correlation) association with liability to EA or to measured EA performance, the opposing effects of alleles from the shared and SZ_diff_ fractions must, more or less, cancel each other out. These findings have important implications for interventions designed to improve educational outcomes in SZ which we suggest may need to focus on noncognitive as well as cognitive mechanisms. They also suggest that there are important shortcomings associated with using EA in genomic studies as a proxy for cognitive function.

Our finding that, in the general population, genetic liability to schizophrenia conferred by common heritable alleles is associated with better educational outcomes than expected given their effects on cognitive ability (Figures 1 and 2) is perhaps surprising given that overall risk of the disorder is associated with poorer EA^64^. Our observation is, however, in keeping with the findings showing that schizophrenia liability is more strongly associated with the extent to which EA in people who develop SZ deviates from that of their family members, and that this deviation is not explained by heritable forms of liability to schizophrenia^65^.

That SZ is more strongly associated than is BD with cognitive impairment led us to postulate that the differentiating liability fraction would be enriched for genes showing high specific expression specificity for prenatal and early childhood developmental stages. However, we did not observe this. This is however in line with the evidence reviewed above that non-familial factors may be more important than common genetic variation in the greater cognitive impairment seen in SZ than BD.

However, to avoid misinterpretation, we stress that the absence of a signal at any developmental stage does not imply genes that are highly expressed during those stages are not enriched for associations, rather it means that genes that characterise those stages by being expressed at higher levels *relative to other developmental stages* are not particularly enriched. Instead, the differentiating fraction was enriched for genes with relatively high expression specificity for early and mid-adulthood, a developmental stage unlikely to substantially influence in the general population either cognition or general educational attainment other than perhaps higher education. This stage of development does, however, correspond to the typical age at onset of psychotic symptoms, the severity of which is associated with the SZ_diff_ fraction in people with bipolar disorder^7^. While our study does not allow causal inferences to be made between enrichment of liability to the differentiating fraction in genes with high expression specificity for any developmental stage and specific symptomatology, we suggest that follow up studies of the genes that are relatively specific to early and mid-adulthood and which are enriched for association to the differentiating fraction might offer a potential window into the biology of psychotic symptomatology. Moreover, given the relevant genes have relatively high expression in adulthood, the implicated biology might be more tractable to therapeutic intervention than those aspects of biology that are more neurodevelopmental.

Our examination of high expression specificity gene-sets to identify candidate cell populations, brain regions, or biological processes, molecular functions, or cellular fractions annotated by gene ontologies did not yield unequivocal associations that might indicate aspects of biology that mediate differential liability to the two disorders. We did find evidence from a mouse brain dataset (sup fig, Supplementary Table S11) that the differentiating fraction was enriched in genes with high expression specificity for striatal medium spiny neurones and for glutamatergic pyramidal cells in the hippocampal CA1 region. This was also true for the shared fraction, although there was very little correlation between the genes responsible between the two gSEM fractions in either cell population. This suggests that while these cell populations may potentially mediate joint and differentiating liability, different genes are responsible for those fractions of liability (Supplementary Table S10).

While of less relevance to the primary aims of the present study, we note that, in contrast to the differentiating signal, the shared signal was enriched in sets of genes with high expression specificity for multiple neuronal populations and gene ontologies. As expected for a shared signal, the same sets were also highlighted by the primary SZ and BD datasets (Supplementary Tables S9-S12, S14), and were related to multiple aspects of neuronal function, development, differentiation, and synaptic transmission, and involved multiple cellular fractions including ion channels, synapses, and both axon and dendritic annotations.

### Strengths and Limitations

We chose to study cognition in a population sample of individuals without severe mental illness to reduce the impact of medication effects and reverse causation. We used both fluid intelligence and a measure of *g*, which we formed from a principal components analysis of four other cognitive tests. We chose these measures based on the available cognitive test data in UK Biobank to ensure that our findings went beyond the analysis of a single cognitive measure and because *g* gives a more robust measure of general cognitive ability^66,67^ and because psychotic disorders are associated with broad, multi-domain cognitive impairments^14^ including in *g*. Moreover, the source GWASs showed associations with cognitive function that were in line with expectations based on the degrees of cognitive impairment seen in the two disorders and seen in previous correlational studies between the disorders and intelligence. This reassures us that the cognitive measures we used were comparable to those used in previous studies demonstrating impaired cognitive function in these disorders. In addition, our results were consistent across the two measures of cognition that we used. A limitation to our interpretation that the discordant findings between effects of liability on measures of cognition and EA point to effects on noncognitive traits that influence EA is that these findings could potentially be explained by aspects of cognition that are not captured by the measures we tapped into to derive *g* or FI. We suggest, however, that our that interpretation is more plausible although there is no way of testing this in the UK Biobank. Individuals in the UK Biobank also differ from individuals in the general population, and in particular they have higher than average levels of educational attainment and cognitive function^23^, which may result in underestimation of the effect sizes of our observed PRS associations to these traits.

## Conclusions

In summary, we have used gSEM to explore the relationships between cognition and shared and differentiating genetic risk for SZ and BD in a large biobank sample with direct measures of both cognition and EA. We find that both the shared and the SZ_diff_ fractions of liability are enriched for alleles that confer risk to poorer cognitive function in the general population, while the BD_diff_ fraction is enriched for alleles associated with better cognition. We also find that despite its negative effects on cognition, the shared fraction is weakly associated with better EA, suggesting the shared fraction influences noncognitive traits that promote better EA, and that these are sufficient to offset the negative cognitive effects of shared liability. In contrast, the SZ differentiating fraction of liability is enriched for alleles that confer risk to poorer EA, and our findings suggest this occurs through both cognitive and non-cognitive mechanisms. We also found evidence that the differentiating fraction was enriched for genes with relatively high expression specificity in early and mid-adulthood, a developmental stage corresponding to the typical age at onset of psychotic symptoms, a clinical feature that has previously been associated with this fraction of liability.

## Data Availability

Genomic SEM data produced available online at the Cardiff Data Repository (https://research-data.cardiff.ac.uk/).

## Notes

### Competing Interest Statement

The authors have declared no competing interest.

### Funding Statement

This study was funded by Innovate UK.

### Author Declarations

PGC Schizophrenia and Bipolar GWAS : https://pgc.unc.edu/for-researchers/download-results/ Skene et al single cell mouse brain expression gene specificity scores: https://doi.org/10.1038/s41588-018-0129-5 Habib et al single nucleus human brain expression gene specificity scores: https://doi.org/10.1038/nmeth.4407 Cameronet al single nucleus human brain expression gene specificity scores: https://doi.org/10.1016/j.biopsych.2022.06.033 Herringsingle nucleus human brain expression gene specificity scores: https://doi.org/10.1016/j.cell.2022.09.039 Brainseq bulk human brain area expression gene developmental stage specificity scores: https://doi.org/10.1016/j.neuron.2019.05.013 ; https://doi.org/10.1016/j.biopsych.2021.03.009

## References

1. Sullivan PF, Daly MJ, O’Donovan M. Genetic architectures of psychiatric disorders: the emerging picture and its implications. Nat Rev Genet. 2012;13(8):537–551. doi:10.1038/nrg3240

2. World Health Organisation. International Statistical Classification of Diseases and Related Health Problems (11th Ed.).; 2019.

3. American Psychiatric Association. Diagnostic and Statistical Manual of Mental Disorders (5th Ed.).; 2013.

4. Craddock N, Owen MJ. The beginning of the end for the Kraepelinian dichotomy. Br J Psychiatry. 2005;186:364–366. doi:10.1192/bjp.186.5.364

5. Brainstorm Consortium, Anttila V, Bulik-Sullivan B, et al. Analysis of shared heritability in common disorders of the brain. Science. 2018;360(6395). doi:10.1126/science.aap8757

6. Owen MJ. New approaches to psychiatric diagnostic classification. Neuron. 2014;84(3):564–571. doi:10.1016/j.neuron.2014.10.028

7. Richards AL, Cardno A, Harold G, et al. Genetic liabilities differentiating bipolar disorder, schizophrenia, and major depressive disorder, and phenotypic heterogeneity in bipolar disorder. JAMA Psychiatry. 2022;79(10):1032–1039. doi:10.1001/jamapsychiatry.2022.2594

8. Allardyce J, Leonenko G, Hamshere M, et al. Association Between Schizophrenia-Related Polygenic Liability and the Occurrence and Level of Mood-Incongruent Psychotic Symptoms in Bipolar Disorder. JAMA Psychiatry. 2018;75(1):28–35. doi:10.1001/jamapsychiatry.2017.3485

9. Bipolar Disorder and Schizophrenia Working Group of the Psychiatric Genomics Consortium. Electronic address, Bipolar Disorder and Schizophrenia Working Group of the Psychiatric Genomics Consortium. Genomic dissection of bipolar disorder and schizophrenia, including 28 subphenotypes. Cell. 2018;173(7):1705–1715.e16. doi:10.1016/j.cell.2018.05.046

10. Dennison CA, Legge SE, Hubbard L, et al. Risk Factors, Clinical Features, and Polygenic Risk Scores in Schizophrenia and Schizoaffective Disorder Depressive-Type. Schizophr Bull. 2021;47(5):1375–1384. doi:10.1093/schbul/sbab036

11. Reichenberg A, Harvey PD, Bowie CR, et al. Neuropsychological function and dysfunction in schizophrenia and psychotic affective disorders. Schizophr Bull. 2009;35(5):1022–1029. doi:10.1093/schbul/sbn044

12. Li W, Zhou F-C, Zhang L, et al. Comparison of cognitive dysfunction between schizophrenia and bipolar disorder patients: A meta-analysis of comparative studies. J Affect Disord. 2020;274:652–661. doi:10.1016/j.jad.2020.04.051

13. Barch DM, Bustillo J, Gaebel W, et al. Logic and justification for dimensional assessment of symptoms and related clinical phenomena in psychosis: relevance to DSM-5. Schizophr Res. 2013;150(1):15–20. doi:10.1016/j.schres.2013.04.027

14. Green MF, Horan WP, Lee J. Nonsocial and social cognition in schizophrenia: current evidence and future directions. World Psychiatry. 2019;18(2):146–161. doi:10.1002/wps.20624

15. Tsitsipa E, Fountoulakis KN. The neurocognitive functioning in bipolar disorder: a systematic review of data. Ann Gen Psychiatry. 2015;14:42. doi:10.1186/s12991-015-0081-z

16. Lynham AJ, Hubbard L, Tansey KE, et al. Examining cognition across the bipolar/schizophrenia diagnostic spectrum. J Psychiatry Neurosci. 2018;43(4):245–253. doi:10.1503/jpn.170076

17. van Haren NEM, Setiaman N, Koevoets MGJC, Baalbergen H, Kahn RS, Hillegers MHJ. Brain structure, IQ, and psychopathology in young offspring of patients with schizophrenia or bipolar disorder. Eur Psychiatry. 2020;63(1):e5. doi:10.1192/j.eurpsy.2019.19

18. Bowie CR, Bell MD, Fiszdon JM, et al. Cognitive remediation for schizophrenia: An expert working group white paper on core techniques. Schizophr Res. 2020;215:49–53. doi:10.1016/j.schres.2019.10.047

19. Craddock N, Owen MJ. The Kraepelinian dichotomy - going, going… but still not gone. Br J Psychiatry. 2010;196(2):92–95. doi:10.1192/bjp.bp.109.073429

20. Owen MJ, O’Donovan MC, Thapar A, Craddock N. Neurodevelopmental hypothesis of schizophrenia. Br J Psychiatry. 2011;198(3):173–175. doi:10.1192/bjp.bp.110.084384

21. Grotzinger AD, Rhemtulla M, de Vlaming R, et al. Genomic structural equation modelling provides insights into the multivariate genetic architecture of complex traits. Nat Hum Behav. 2019;3(5):513–525. doi:10.1038/s41562-019-0566-x

22. Power RA, Steinberg S, Bjornsdottir G, et al. Polygenic risk scores for schizophrenia and bipolar disorder predict creativity. Nat Neurosci. 2015;18(7):953–955. doi:10.1038/nn.4040

23. Hagenaars SP, Harris SE, Davies G, et al. Shared genetic aetiology between cognitive functions and physical and mental health in UK Biobank (N=112 151) and 24 GWAS consortia. Mol Psychiatry. 2016;21(11):1624–1632. doi:10.1038/mp.2015.225

24. Hill WD, Marioni RE, Maghzian O, et al. A combined analysis of genetically correlated traits identifies 187 loci and a role for neurogenesis and myelination in intelligence. Mol Psychiatry. 2019;24(2):169–181. doi:10.1038/s41380-017-0001-5

25. Okbay A, Beauchamp JP, Fontana MA, et al. Genome-wide association study identifies 74 loci associated with educational attainment. Nature. 2016;533(7604):539-542. doi:10.1038/nature17671

26. Escott-Price V, Bracher-Smith M, Menzies G, et al. Genetic liability to schizophrenia is negatively associated with educational attainment in UK Biobank. Mol Psychiatry. 2020;25(4):703–705. doi:10.1038/s41380-018-0328-6

27. Hindley G, Frei O, Shadrin AA, et al. Charting the landscape of genetic overlap between mental disorders and related traits beyond genetic correlation. Am J Psychiatry. 2022;179(11):833–843. doi:10.1176/appi.ajp.21101051

28. Shafee R, Nanda P, Padmanabhan JL, et al. Polygenic risk for schizophrenia and measured domains of cognition in individuals with psychosis and controls. Transl Psychiatry. 2018;8(1):78. doi:10.1038/s41398-018-0124-8

29. Owen MJ, O’Donovan MC. The genetics of cognition in schizophrenia. Genomic Psychiatry. July 16, 2024:1–8. doi:10.61373/gp024i.0040

30. Mullins N, Forstner AJ, O’Connell KS, et al. Genome-wide association study of more than 40,000 bipolar disorder cases provides new insights into the underlying biology. Nat Genet. 2021;53(6):817–829. doi:10.1038/s41588-021-00857-4

31. Trubetskoy V, Pardiñas AF, Qi T, et al. Mapping genomic loci implicates genes and synaptic biology in schizophrenia. Nature. 2022;604(7906):502-508. doi:10.1038/s41586-022-04434-5

32. International HapMap 3 Consortium, Altshuler DM, Gibbs RA, et al. Integrating common and rare genetic variation in diverse human populations. Nature. 2010;467(7311):52–58. doi:10.1038/nature09298

33. Bulik-Sullivan B, Loh P-R, Finucane HK, et al. LD Score regression distinguishes confounding from polygenicity in genome-wide association studies. Nat Genet. 2015;47(3):291–295. doi:10.1038/ng.3211

34. Bulik-Sullivan B, Finucane HK, Anttila V, et al. An atlas of genetic correlations across human diseases and traits. Nat Genet. 2015;47(11):1236–1241. doi:10.1038/ng.3406

35. Savage JE, Jansen PR, Stringer S, et al. Genome-wide association meta-analysis in 269,867 individuals identifies new genetic and functional links to intelligence. Nat Genet. 2018;50(7):912–919. doi:10.1038/s41588-018-0152-6

36. Okbay A, Wu Y, Wang N, et al. Polygenic prediction of educational attainment within and between families from genome-wide association analyses in 3 million individuals. Nat Genet. 2022;54(4):437–449. doi:10.1038/s41588-022-01016-z

37. International Schizophrenia Consortium, Purcell SM, Wray NR, et al. Common polygenic variation contributes to risk of schizophrenia and bipolar disorder. Nature. 2009;460(7256):748–752. doi:10.1038/nature08185

38. Legge SE, Pardiñas AF, Woolway G, et al. Genetic and phenotypic features of schizophrenia in the UK biobank. JAMA Psychiatry. 2024;81(7):681–690. doi:10.1001/jamapsychiatry.2024.0200

39. Fawns-Ritchie C, Deary IJ. Reliability and validity of the UK Biobank cognitive tests. PLoS ONE. 2020;15(4):e0231627. doi:10.1371/journal.pone.0231627

40. Lyall DM, Cullen B, Allerhand M, et al. Cognitive test scores in UK biobank: data reduction in 480,416 participants and longitudinal stability in 20,346 participants. PLoS ONE. 2016;11(4):e0154222. doi:10.1371/journal.pone.0154222

41. Spearman C. “general intelligence” objectively determined and measured. In: Jenkins JJ, Paterson DG, eds. Studies in Individual Differences: The Search for Intelligence. Appleton-Century-Crofts; 1961:59–73. doi:10.1037/11491-006

42. Fenner E, Holmans P, O’Donovan MC, Owen MJ, Walters JT, Rees E. Rare coding variants in schizophrenia-associated genes affect generalised cognition in the UK Biobank. medRxiv. August 16, 2023. doi:10.1101/2023.08.14.23294074

43. Leonenko G, Di Florio A, Allardyce J, et al. A data-driven investigation of relationships between bipolar psychotic symptoms and schizophrenia genome-wide significant genetic loci. Am J Med Genet B Neuropsychiatr Genet. 2018;177(4):468–475. doi:10.1002/ajmg.b.32635

44. Cameron D, Mi D, Vinh N-N, et al. Single-Nuclei RNA Sequencing of 5 Regions of the Human Prenatal Brain Implicates Developing Neuron Populations in Genetic Risk for Schizophrenia. Biol Psychiatry. 2023;93(2):157–166. doi:10.1016/j.biopsych.2022.06.033

45. Herring CA, Simmons RK, Freytag S, et al. Human prefrontal cortex gene regulatory dynamics from gestation to adulthood at single-cell resolution. Cell. 2022;185(23):4428–4447.e28. doi:10.1016/j.cell.2022.09.039

46. Tume CE, Chick SL, Holmans PA, et al. Genetic implication of specific glutamatergic neurons of the prefrontal cortex in the pathophysiology of schizophrenia. Biological Psychiatry Global Open Science. 2024;4(5):100345. doi:10.1016/j.bpsgos.2024.100345

47. Skene NG, Bryois J, Bakken TE, et al. Genetic identification of brain cell types underlying schizophrenia. Nat Genet. 2018;50(6):825–833. doi:10.1038/s41588-018-0129-5

48. Habib N, Avraham-Davidi I, Basu A, et al. Massively parallel single-nucleus RNA-seq with DroNc-seq. Nat Methods. 2017;14(10):955–958. doi:10.1038/nmeth.4407

49. de Leeuw CA, Mooij JM, Heskes T, Posthuma D. MAGMA: generalized gene-set analysis of GWAS data. PLoS Comput Biol. 2015;11(4):e1004219. doi:10.1371/journal.pcbi.1004219

50. Finucane HK, Reshef YA, Anttila V, et al. Heritability enrichment of specifically expressed genes identifies disease-relevant tissues and cell types. Nat Genet. 2018;50(4):621–629. doi:10.1038/s41588-018-0081-4

51. Collado-Torres L, Burke EE, Peterson A, et al. Regional Heterogeneity in Gene Expression, Regulation, and Coherence in the Frontal Cortex and Hippocampus across Development and Schizophrenia. Neuron. 2019;103(2):203–216.e8. doi:10.1016/j.neuron.2019.05.013

52. Robinson MD, McCarthy DJ, Smyth GK. edgeR: a Bioconductor package for differential expression analysis of digital gene expression data. Bioinformatics. 2010;26(1):139–140. doi:10.1093/bioinformatics/btp616

53. Clifton NE, Hannon E, Harwood JC, et al. Dynamic expression of genes associated with schizophrenia and bipolar disorder across development. Transl Psychiatry. 2019;9(1):74. doi:10.1038/s41398-019-0405-x

54. Clifton NE, Collado-Torres L, Burke EE, et al. Developmental Profile of Psychiatric Risk Associated With Voltage-Gated Cation Channel Activity. Biol Psychiatry. 2021;90(6):399–408. doi:10.1016/j.biopsych.2021.03.009

55. Ashburner M, Ball CA, Blake JA, et al. Gene Ontology: Tool for the unification of biology. Nat Genet. 2000;25(1):25–29. doi:10.1038/75556

56. Gene Ontology Consortium, Aleksander SA, Balhoff J, et al. The Gene Ontology knowledgebase in 2023. Genetics. 2023;224(1):iyad031. doi:10.1093/genetics/iyad031

57. Sniekers S, Stringer S, Watanabe K, et al. Genome-wide association meta-analysis of 78,308 individuals identifies new loci and genes influencing human intelligence. Nat Genet. 2017;49(7):1107–1112. doi:10.1038/ng.3869

58. Murray RM, Sham P, Van Os J, Zanelli J, Cannon M, McDonald C. A developmental model for similarities and dissimilarities between schizophrenia and bipolar disorder. Schizophr Res. 2004;71(2-3):405–416. doi:10.1016/j.schres.2004.03.002

59. Kendler KS, Ohlsson H, Keefe RSE, Sundquist K, Sundquist J. The joint impact of cognitive performance in adolescence and familial cognitive aptitude on risk for major psychiatric disorders: a delineation of four potential pathways to illness. Mol Psychiatry. 2018;23(4):1076–1083. doi:10.1038/mp.2017.78

60. Smeland OB, Bahrami S, Frei O, et al. Genome-wide analysis reveals extensive genetic overlap between schizophrenia, bipolar disorder, and intelligence. Mol Psychiatry. 2020;25(4):844–853. doi:10.1038/s41380-018-0332-x

61. Werme J, van der Sluis S, Posthuma D, de Leeuw CA. An integrated framework for local genetic correlation analysis. Nat Genet. 2022;54(3):274–282. doi:10.1038/s41588-022-01017-y

62. Nieuwboer HA, Pool R, Dolan CV, Boomsma DI, Nivard MG. GWIS: Genome-Wide Inferred Statistics for Functions of Multiple Phenotypes. Am J Hum Genet. 2016;99(4):917–927. doi:10.1016/j.ajhg.2016.07.020

63. Bansal V, Mitjans M, Burik CAP, et al. Genome-wide association study results for educational attainment aid in identifying genetic heterogeneity of schizophrenia. Nat Commun. 2018;9(1):3078. doi:10.1038/s41467-018-05510-z

64. Kendler KS, Ohlsson H, Mezuk B, Sundquist K, Sundquist J. A Swedish National Prospective and Co-relative Study of School Achievement at Age 16, and Risk for Schizophrenia, Other Nonaffective Psychosis, and Bipolar Illness. Schizophr Bull. 2016;42(1):77–86. doi:10.1093/schbul/sbv103

65. Kendler KS, Ohlsson H, Mezuk B, Sundquist JO, Sundquist K. Observed cognitive performance and deviation from familial cognitive aptitude at age 16 years and ages 18 to 20 years and risk for schizophrenia and bipolar illness in a swedish national sample. JAMA Psychiatry. 2016;73(5):465–471. doi:10.1001/jamapsychiatry.2016.0053

66. Spearman C. “General Intelligence,” Objectively Determined and Measured. Am J Psychol. 1904;15(2):201. doi:10.2307/1412107

67. Cattell RB. The measurement of adult intelligence. Psychol Bull. 1943;40(3):153–193. doi:10.1037/h0059973

