## Supplementary Figures S1 to S6 for "The relationship between shared and differentiating genetic liability for schizophrenia and bipolar disorder and cognition and educational attainment in the UK Biobank"

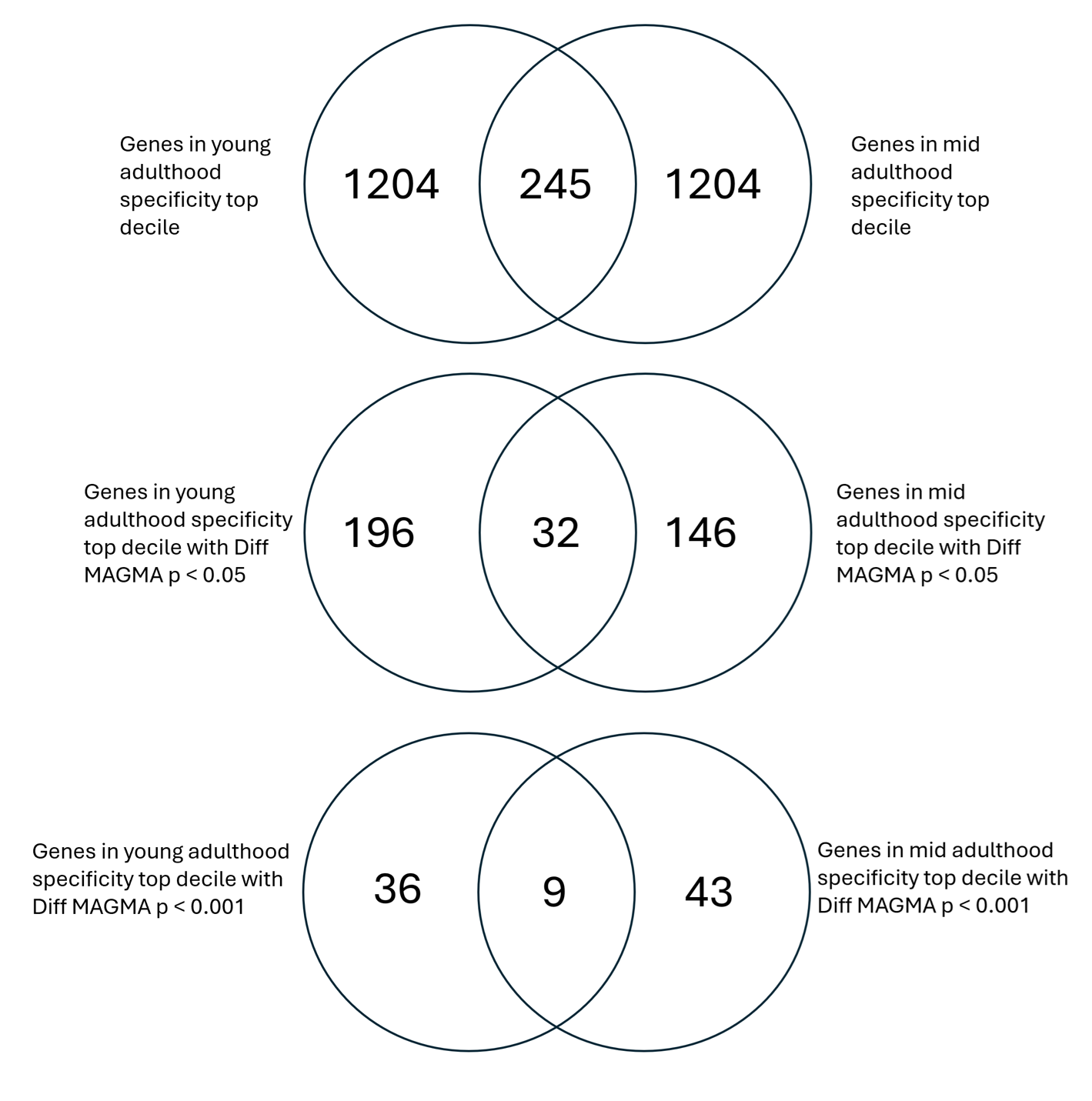

Supplementary Figure 1. Venn diagrams showing overlap between genes in top deciles of specificity for young adulthood and mid adulthood

**Habib *et al***

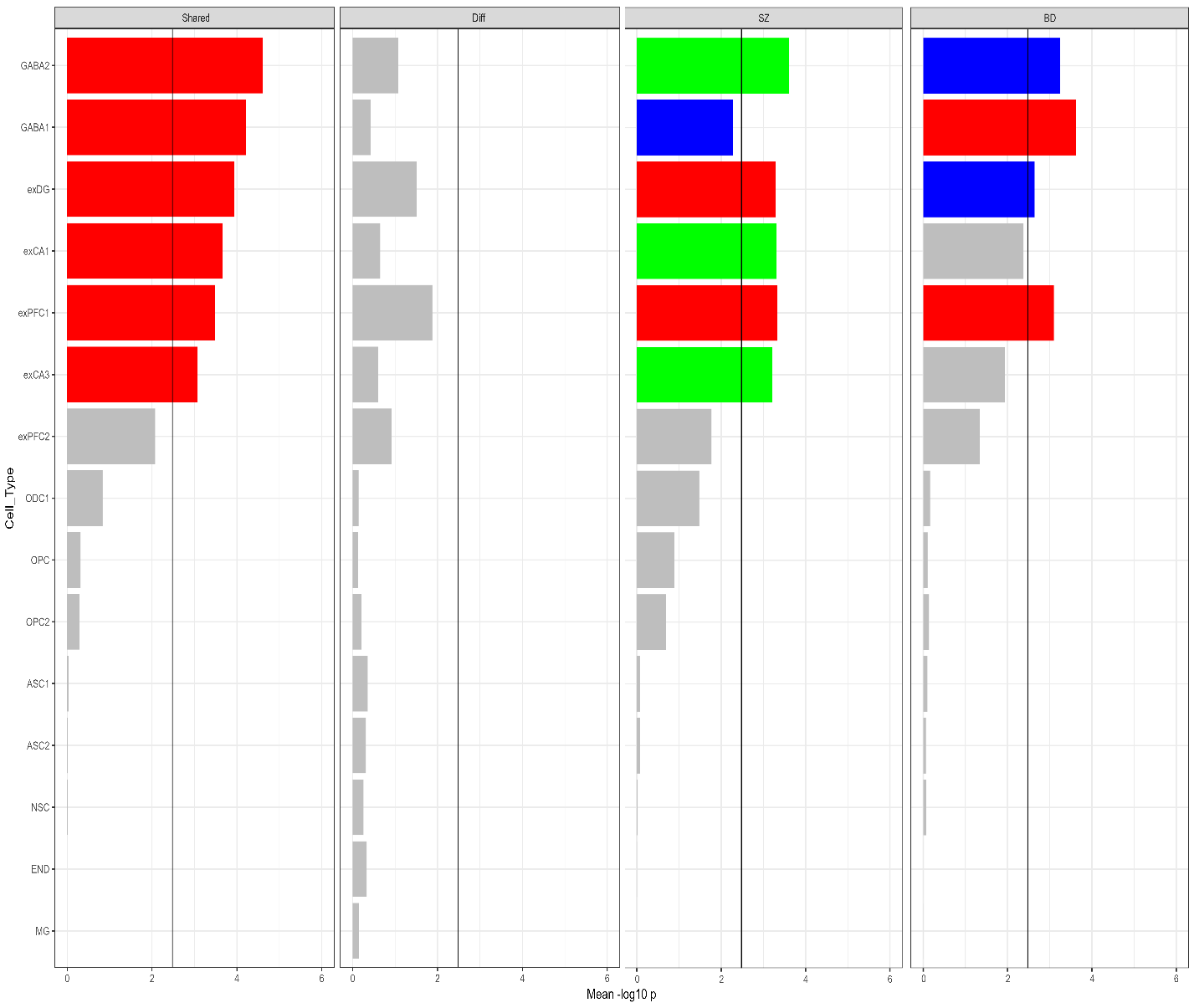

Figure S2. Enrichment of genomic SEM SZ/BD shared and differentiating components. Gene sets tested for enrichment comprise genes with the top 10% expression specificity values for the cell types in the Habib adult hippocampus and prefrontal cortex single nucleus RNA sequencing dataset[^1^](https://sciwheel.com/work/citation?ids=4114189&pre=&suf=&sa=0&dbf=0). Mean -log10 p indicates mean -log significance level across two enrichment analysis methods, MAGMA and partitioned LDSC. Black line represents significance threshold (Bonferroni corrected for 15 cell types). Red bars indicate significance threshold reached in both MAGMA and pLDSC, green in MAGMA only, and blue in pLDSC only.

**Skene *et al***

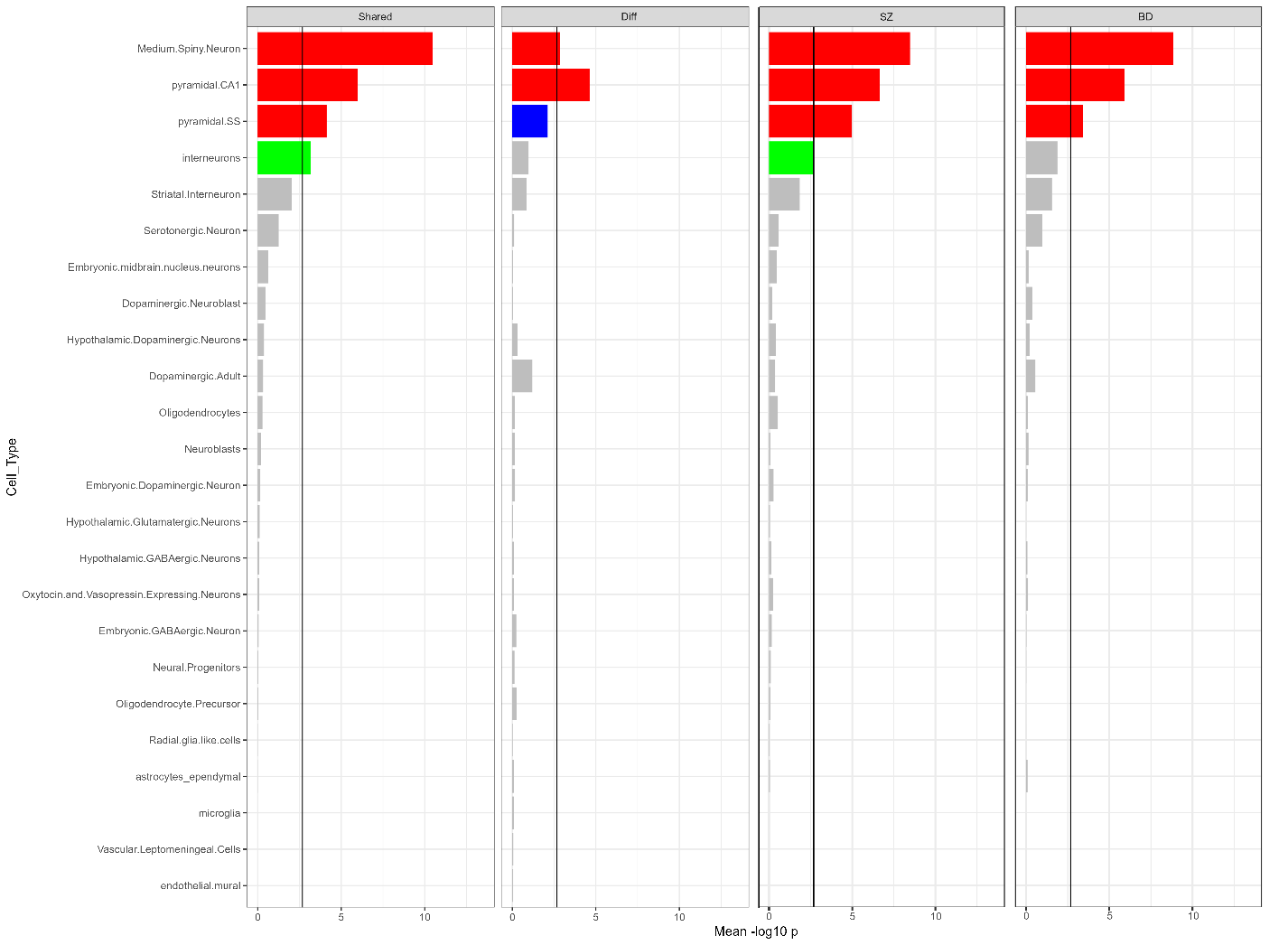

Figure S3. Enrichment of genomic SEM SZ/BD shared and differentiating components. Gene sets tested for enrichment comprise genes with the top 10% expression specificity values for the cell types in the Skene adult and fetal brain single nucleus RNA sequencing dataset[^2^](https://sciwheel.com/work/citation?ids=5281403&pre=&suf=&sa=0&dbf=0). Mean -log10 p indicates mean -log significance level across two enrichment analysis methods, MAGMA and partitioned LDSC. Black line represents significance threshold (Bonferroni corrected for 24 cell types). Red bars indicate significance threshold reached in both MAGMA and pLDSC, green in MAGMA only, and blue in pLDSC only.

**Herring *et al***

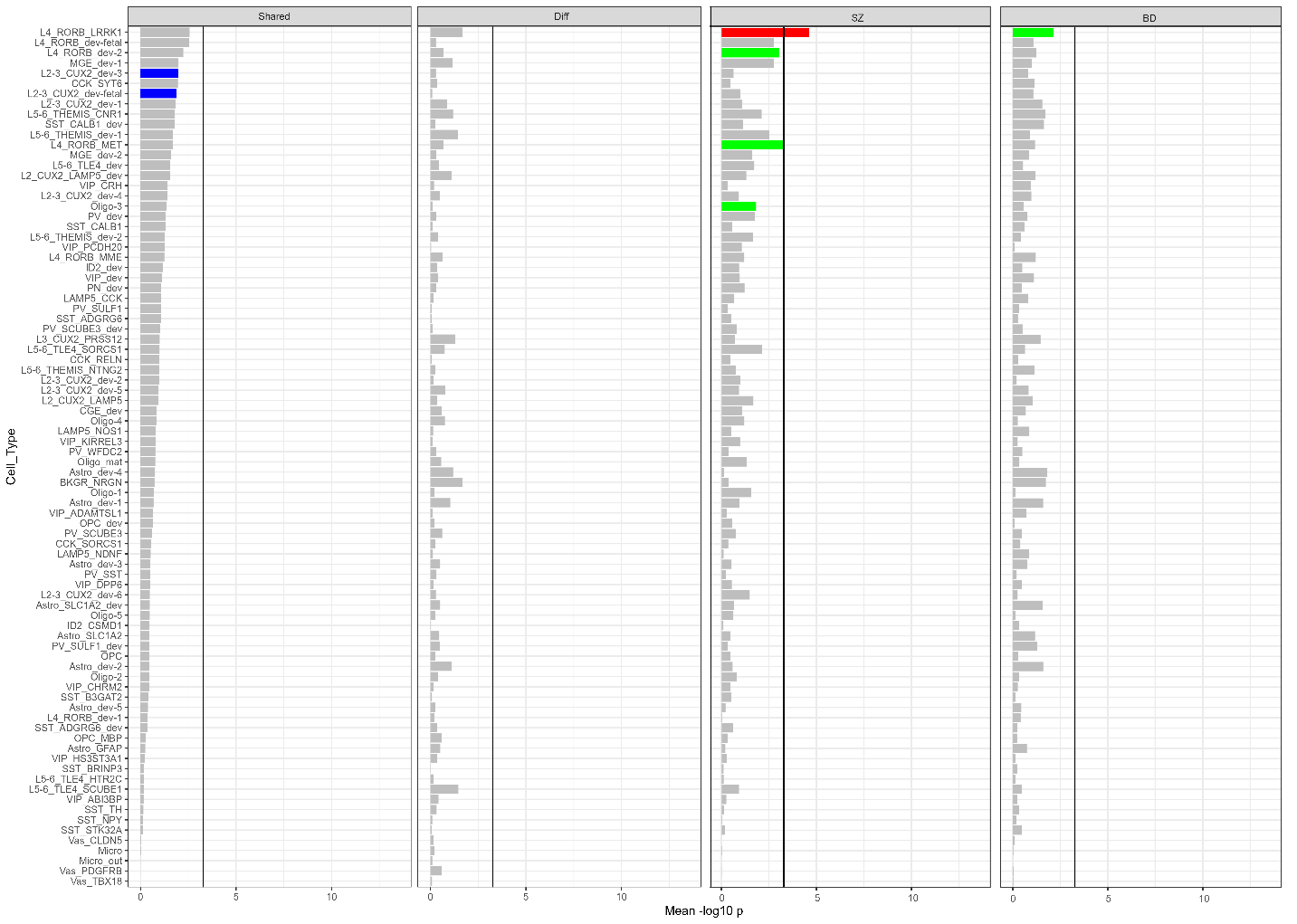

Figure S4. Enrichment of genomic SEM SZ/BD shared and differentiating components. Gene sets tested for enrichment comprise genes with the top 10% expression specificity values for the cell types in the Herring fetal, neonatal and adult prefrontal cortex single nucleus RNA sequencing dataset[^3^](https://sciwheel.com/work/citation?ids=13865003&pre=&suf=&sa=0&dbf=0). Mean -log10 p indicates mean -log significance level across two enrichment analysis methods, MAGMA and partitioned LDSC. Black line represents significance threshold (Bonferroni corrected for 84 cell types). Red bars indicate significance threshold reached in both MAGMA and pLDSC, green in MAGMA only, and blue in pLDSC only.

**Cameron, Bray *et al***

All Cameron *et al* results divided across two graphs for readability.

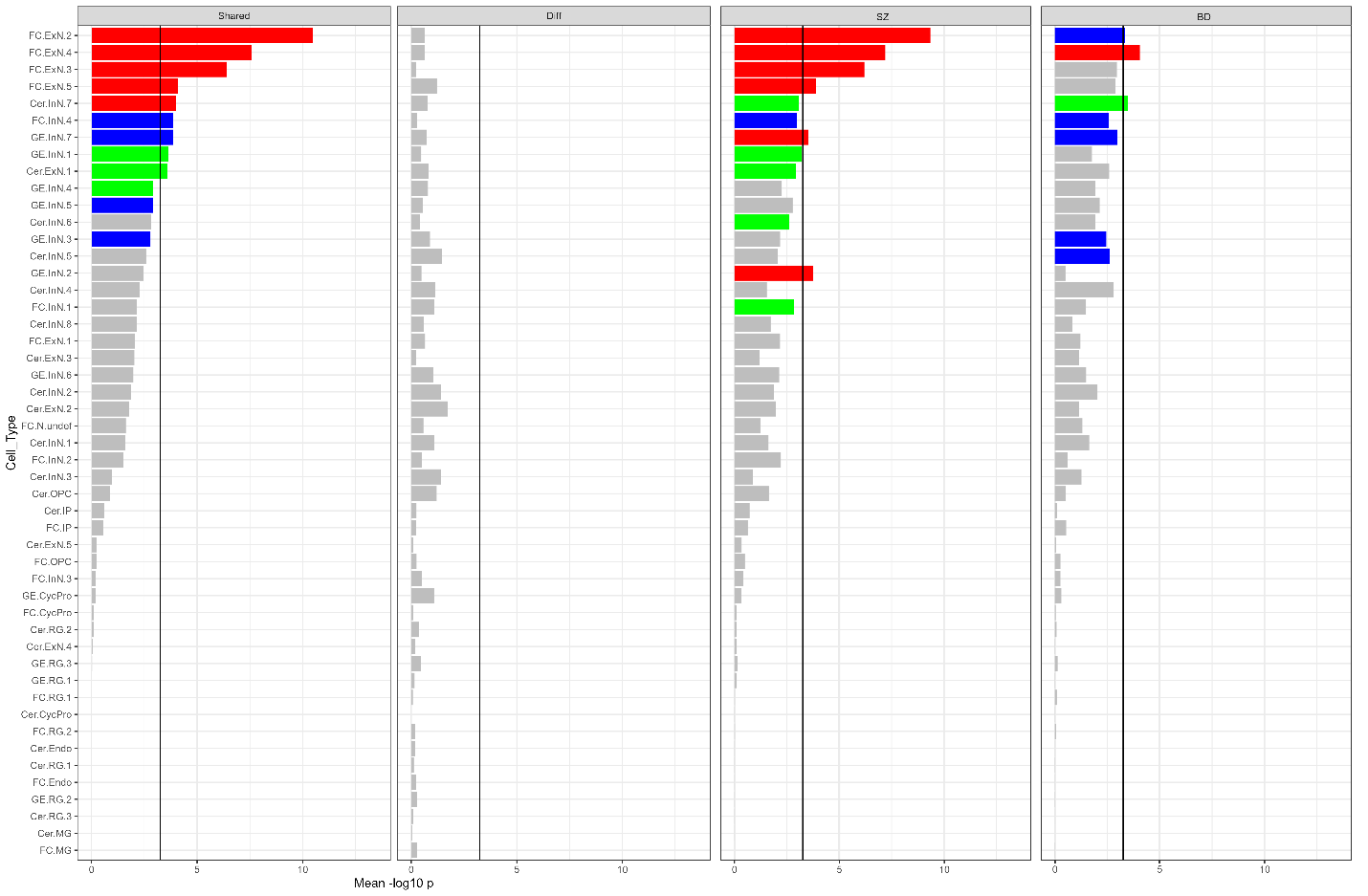

Figure S5. Enrichment of genomic SEM SZ/BD shared and differentiating components. Gene sets tested for enrichment comprise genes with the top 10% expression specificity values for the cell types in the Cameron second trimester fetal brain single nucleus RNA sequencing dataset (frontal cortex, ganglionic eminence and cerebellum shown in this figure)[^4^](https://sciwheel.com/work/citation?ids=13704448&pre=&suf=&sa=0&dbf=0). Mean -log10 p indicates mean -log significance level across two enrichment analysis methods, MAGMA and partitioned LDSC. Black line represents significance threshold (Bonferroni corrected for 91 cell types across all five brain areas). Red bars indicate significance threshold reached in both MAGMA and pLDSC, green in MAGMA only, and blue in pLDSC only.

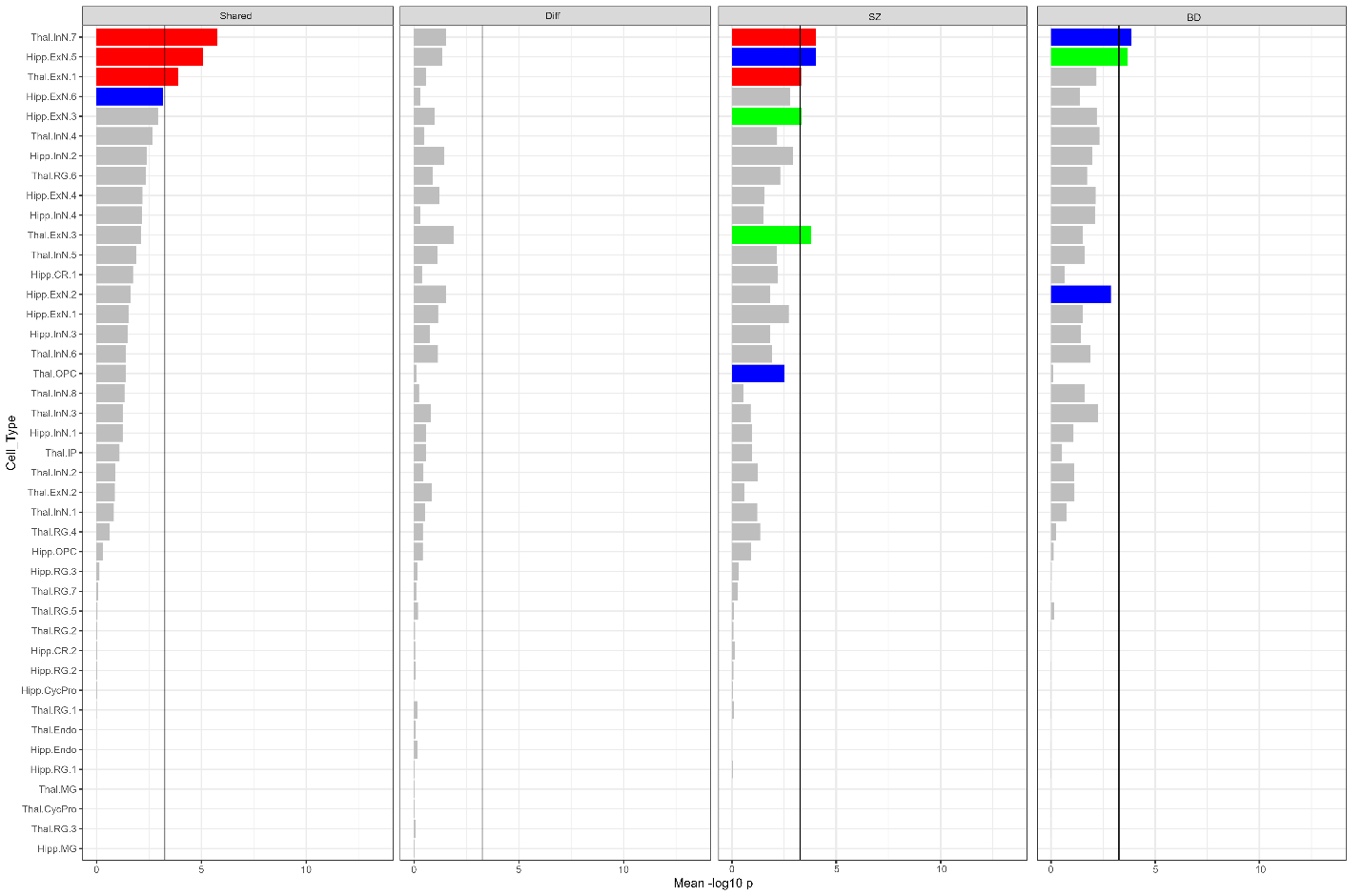

Figure S6. Enrichment of genomic SEM SZ/BD shared and differentiating components. Gene sets tested for enrichment comprise genes with the top 10% expression specificity values for the cell types in the Cameron second trimester fetal brain single nucleus RNA sequencing dataset (hippocampus and thalamus shown in this figure)[^4^](https://sciwheel.com/work/citation?ids=13704448&pre=&suf=&sa=0&dbf=0). Mean -log10 p indicates mean -log significance level across two enrichment analysis methods, MAGMA and partitioned LDSC. Black line represents significance threshold (Bonferroni corrected for 91 cell types across all five brain areas). Red bars indicate significance threshold reached in both MAGMA and pLDSC, green in MAGMA only, and blue in pLDSC only.

[1.    Habib N, Avraham-Davidi I, Basu A, et al. Massively parallel single-nucleus RNA-seq with DroNc-seq. *Nat Methods*. 2017;14(10):955-958. doi:10.1038/nmeth.4407](https://sciwheel.com/work/bibliography/4114189)

[2.    Skene NG, Bryois J, Bakken TE, et al. Genetic identification of brain cell types underlying schizophrenia. *Nat Genet*. 2018;50(6):825-833. doi:10.1038/s41588-018-0129-5](https://sciwheel.com/work/bibliography/5281403)

[3.    Herring CA, Simmons RK, Freytag S, et al. Human prefrontal cortex gene regulatory dynamics from gestation to adulthood at single-cell resolution. *Cell*. 2022;185(23):4428-4447.e28. doi:10.1016/j.cell.2022.09.039](https://sciwheel.com/work/bibliography/13865003)

[4.    Cameron D, Mi D, Vinh N-N, et al. Single-Nuclei RNA Sequencing of 5 Regions of the Human Prenatal Brain Implicates Developing Neuron Populations in Genetic Risk for Schizophrenia. *Biol Psychiatry*. 2023;93(2):157-166. doi:10.1016/j.biopsych.2022.06.033](https://sciwheel.com/work/bibliography/13704448)
