## Supplementary Methods for "The relationship between shared and differentiating genetic liability for schizophrenia and bipolar disorder and cognition and educational attainment in the UK Biobank"

**lavaan code for two models**

Shared model

F1 =~ a*SZ + a*BD

F1 ~ SNP

F1 ~~ 1*F1

This model creates a shared fraction (F1) and examines its effect on each SNP. The number of parameters, including the output of interest, that gSEM can estimate is limited by the number of inputs, here the SZ GWAS and BD GWAS. To make the model identifiable and possible to run, it is necessary to constrain the number of parameters the model needs to estimate. The model code achieves this by loading each source GWAS onto the shared component equally (line 1 of code) and constraining the variance of the shared fraction to 1 (line 3 of code).

Differentiating model

F1 =~ SZ + BD

F2 =~ SZ

F1 ~ SNP

F2 ~ SNP

F1 ~~ F1

F2 ~~ F2

F1 ~~ 0*F2

SZ ~~ 0*SZ

BD ~~ 0*BD

SZ ~~ 0*BD

This model creates a shared fraction (F1) from the SZ and BD source GWAS, and an F2 fraction that captures the residual variance not captured by the shared component (lines 1 and 2). The effect of these fractions on each SNP are assessed (lines 3 and 4). The F1 and F2 components have their variance constrained to 1 to make the model identifiable (lines 5 and 6). They are minimally correlated with each other (line 7), and have their residual variance and covariance set to 0 (lines 8-10).
